# Investigating the Effects of Artificial Baroreflex Stimulation on Pain Perception: A Comparative Study in Healthy Participants and Individuals with Chronic Low Back Pain

**DOI:** 10.1101/2023.12.18.23299896

**Authors:** Alessandra Venezia, Harriet-Fawsitt Jones, David Hohenschurz-Schmidt, Matteo Mancini, Matthew Howard, Elena Makovac

**Affiliations:** Department of Neuroimaging, Institute of Psychology, Psychiatry & Neuroscience, King’s College London, London, UK; Department of Surgery & Cancer, Imperial College London, London, UK; Department of Cardiovascular, Endocrine-Metabolic Diseases and Aging, Italian National Institute of Health, Rome, Italy; Department of Life Sciences, Division of Psychology, Brunel University London, London, UK

**Keywords:** Baroreceptor Sensitivity Index, Artificial Baroreceptor stimulation, Chronic Lower Back Pain, Conditioned Pain Modulation, Pressure Pain

## Abstract

The autonomic nervous system (ANS) and pain exhibit a reciprocal relationship, whereupon acute pain triggers ANS responses, while resting ANS activity can influence pain perception. Nociceptive signalling can also be altered by “top-down” processes occurring in the brain, brainstem, and spinal cord, known as *descending modulation*. By employing the Conditioned Pain Modulation (CPM) paradigm, our previous study revealed a connection between reduced low-frequency heart rate variability (HRV) and CPM. Chronic pain patients often experience both ANS dysregulation and impaired CPM. Baroreceptors, which contribute to blood pressure and HRV regulation, may play a significant role in this relationship, but their involvement in pain perception and their functioning in chronic pain have not been sufficiently explored. In this study, we combined artificial *baroreceptor stimulation* in both pressure pain and CPM paradigms, seeking to explore the role of baroreceptors in pain perception and descending modulation. 22 patients with chronic low back pain (CLBP) and 29 healthy controls (HC) took part in this study. We identified a relationship between baroreflex functioning and perception of pressure pain, finding differential modulation of pressure pain between diagnostic groups. Specifically, HC participants perceived less pain in response to baroreflex activation, whereas CLBP patients exhibited increased pain sensitivity. CPM scores were associated with baseline measures of baroreflex efficiency in both patients and controls. Our data support the importance of the baroreflex in chronic pain and a possible mechanism of dysregulation involving the interaction between the autonomic nervous system and descending pain modulation.

## 1. Introduction

The interplay between the cardiovascular system and pain plays a crucial role in pain regulation. In healthy controls (HC), acute pain increases sympathetic arousal and blood pressure (BP)(1). Conversely, a reduction in pain perception is reported in HC during spontaneous (2) or induced (3) high BP, as well as in unmedicated patients with hypertension (4). Patients receiving angiotensin-converting enzyme inhibitors (commonly used to treat hypertension) are more likely to have chronic pain, compared to untreated individuals (5). In sum, several studies have suggested that high BP may protect against clinical pain (6), highlighting the existence of a homeostatic regulative mechanism of pain perception.

Nociception is also modulated by ‘top-down’ processes in brain, brainstem and spinal cord. Descending pain modulation (7, 8) can be triggered by a tonic painful conditioning stimulus, which can suppress incoming nociceptive signals arising from a second acute stimulus at a different body site. Conditioned Pain Modulation (CPM) is a widely used paradigm for assessing descending pain modulation in humans (9). A deficient descending pain modulation, as measured by CPM, has been suggested to be one of the mechanisms involved in the persistence of pain. Reduced CPM efficiency has been demonstrated in various chronic pain states, including fibromyalgia (see (10) for a meta-analysis on the topic), chronic migraine (11), and chronic temporomandibular pain disorder (12). A recent meta-analysis in chronic low back pain (CLBP) patients, however, showed that CPM efficiency in this cohort is controversial;, three published studies found significant differences in CPM between CLBP and HC, whilst four studies did not find a difference (13). These discrepancies might be the consequence of methodological choices (14) or biopsychosocial factors, including pain catastrophizing, gender, age, and beliefs (15, 16). Interestingly, heterogeneity in pathophysiological mechanisms related to CPM has been described, indicating that chronic pain patients can be clustered into different groups based on their CPM profile (17).

An association has been also described between BP and CPM response (18). The stimulus used in the CPM paradigm elicits pain, but also an accompanying cardiovascular response. Here, baroreceptors may play a central role, signalling the cardiovascular state in the body to the brain. Artificial baroreceptor stimulation results in reduced pain sensitivity in both hypertensive and normotensive individuals (19). Baroreceptor stimulation has been also shown to attenuate attentional effects to pain stimuli (20). Some pioneering studies have suggested that the pain-attenuating effect of baroreflex stimulation is disrupted in chronic pain (21), arguing that efficient descending pain modulation depends on effective autonomic regulation and that deficiencies in these mechanisms relates to pain persistence. Chronic pain patients exhibit both autonomic dysregulation (22) and deficient CPM (23), suggesting a causal relationship between a dysregulated ANS, deficient descending pain modulation, and pain chronicity, possibly mediated by a dysfunction in the baroreflex.

Here, we aimed to examine the relationship between ANS reactivity and CPM efficiency in HC and patients with CLBP, with a focus on the specific involvement of baroreceptors in this interaction. We examined the impact of autonomic perturbation on pain perception using artificial baroreceptor stimulation during a pressure pain paradigm. We also assessed the efficiency of descending pain modulation (CPM) in changing pain sensations. We hypothesized that the efficiency of the baroreflex would be linked with pain perception and modulation in HC, but that this association would be disrupted in CLBP patients.

## Methods

### 2.1 Ethical approval

The study was approved by King’s College London Research Ethics Committee (HR-19/20-14149) and was conducted in accordance with the Declaration of Helsinki. All participants were fully informed regarding the experimental procedures and provided written informed consent prior to participating.

### 2.2 Participants

Twenty-two CLBP patients and twenty-nine age- and gender-matched healthy control participants (HC) took part in this study. All participants were right-handed. CLBP was defined as continuous or recurrent episodes of pain in the lower back (with or without pain in a lower extremity) that persisted or recurred over the past 3 months (24, 25). Other inclusion criteria for patients were: (1) age between 18 and 65 years; (2) no structural pathology, and (3) under stable or no pharmacological management.

Exclusion criteria included a history of brain injuries, hypertension, neurological or psychiatric disease, and alcohol or drug abuse. Additional cardiovascular exclusion criteria included: personal or family history of hypertension; smoking more than 5 cigarettes a day; BMI > 30; a history of orthostatic hypotension or carotid hypersensitivity. At the beginning of each visit, participants were tested for drug use and alcohol consumption using a urine drug screen and alcohol breathalyser test, respectively. To further exclude possible alterations of the ANS, a Valsalva Manoeuvre was performed at the beginning of each session, whereby a decrease in systolic pressure of > 20 mmHg was defined as abnormal ANS functioning and specified as an exclusion criterion. Prior to each session, participants were required to abstain from alcohol for 24 hours, non-steroidal anti-inflammatory drugs and paracetamol for 12 hours, tobacco and nicotine-containing products for 4 hours, and to limit caffeine intake to a maximum of one caffeinated drink.

### 2.3 Experimental setup for *baroreceptor stimulation*

A bespoke baroreceptor stimulating device was developed by engineers at the Department of Neuroimaging at KCL. The device used two individual cuffs to apply non-painful negative pressure (−60 mmHg) for efficacious baroreceptor stimulation (ACTIVE condition) and −20mmHg stimulation (SHAM condition) applied to the neck at the location of the carotid bifurcation. The device has been used safely in previous HC studies (26–28) to deliver reliable modulation of peripheral (cardiovascular) and central (neuronal) mechanisms (26, 27).

### 2.4 General overview of the experimental procedures

Participants took part in one experimental session which lasted approximately two hours (Figure 1). At the beginning of each experimental session, participants were asked to rate the intensity level of their current pain (Pain State) and their average pain levels during the past year (Pain Trait, (29)), using a Numerical Rating scale (NRS) from 0-100 (0 indicating no pain, 100 indicating worse imaginable pain).

**Figure 1.**
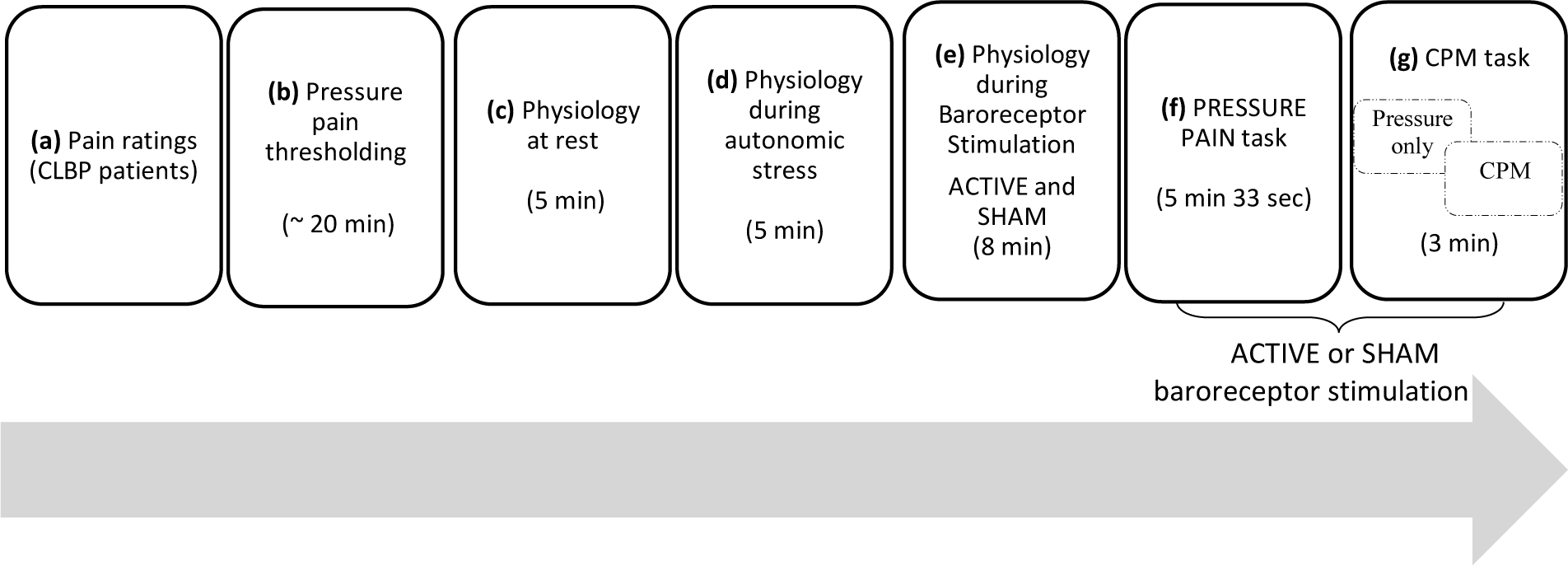
Graphical representation of the experimental session. **(a)**Participants rated their current pain and their average pain during the past year. **(b)** Pain thresholding-individual pressure pain thresholds and a moderate pain level corresponding to a 70/100 visual analogue scale (VAS) were determined. **(c-d)** Baseline (resting) physiology and physiology in response to an autonomic stress (cold pain), each measured for 5 minutes**. (e)** Physiology measurement during ACTIVE and SHAM baroreceptor stimulation. **(f-g)** PRESSURE PAIN and the CPM tasks. Participants rated the intensity of pressure pain presented on the right thumbnail (PRESSURE PAIN task) or simultaneously on the left and right thumbnail (CPM task). In both (f) and (g) pressure pain stimuli were presented coincident with either ACTIVE or SHAM baroreceptor stimulation.

Next, a pain thresholding session was conducted to identify participants’ individual pressure pain thresholds. For a thorough characterisation of the baroreflex activity, participants’ ANS activity was tested at rest (REST condition), during autonomic stress induced by a tonic cold pain stimulus (STRESS condition), or in response to baroreceptor stimulation (ACTIVE condition). Finally, participants completed two experimental tasks, both of which involved pressure stimulation. First, a pain task, during which noxious pressure was delivered coincident with simultaneous ACTIVE or SHAM baroreceptor stimulation. The second was a CPM experiment, in which pressure stimulation was delivered to participants’ right thumbnails, under two conditions; a test stimulus- (‘Pressure only’ baseline condition), or with simultaneous bilateral pressure stimulation (‘CPM’ condition), During the CPM condition, pressure stimuli were delivered to the right thumbnail (test stimulus)- and left thumbnail (conditioning stimulus). These stimuli were both presented with either ACTIVE or SHAM baroreceptor stimulation.

#### 2.4.1 Pain ratings (Figure 1, a)

Following recent publications highlighting the importance of measuring stable and transitory pain levels in chronic pain patients (29), CLBP patients were asked to rate their usual levels of low back pain (Pain Trait) and their current levels of pain (Pain State). Pain Trait was defined as the average intensity of low back pain experienced over the past year, rated on a scale ranging from 0 (no pain) to 100 (the most severe imaginable pain). Pain State, on the other hand, referred to the specific level of lower back pain experienced on the day of the experimental testing, also measured on a 0-100 scale.

#### 2.4.2 Pressure pain thresholding (Figure 1, b)

Sensory thresholding for pressure pain was performed using an established ascending sequential staircase paradigm (30). We used a custom-designed pain pressure probe, which provided perpendicular force to the thumbnail. The ascending staircase paradigm commenced with an initial force of 23 kilopascals (kPa) applied to the nail surface for 2 seconds. Incremental increases of 3.3 kPa were subsequently applied. Participants indicated when they first felt pain (their pain detection threshold), and when they reached a moderate-to-severe level of pain, corresponding to 70/100. The maximum force that could be applied was 138 kPa. Next, participants received 15 pressure stimulations, delivered in a pseudo-random order. The minimum pressure delivered corresponded to each participant’s pain threshold and the maximum pressure to their 70/100 rating. Three intermediate pressure pain levels were also delivered, each equally spaced from one another. Participants rated each stimulation immediately afterward. Participants’ final 70/100 threshold was then calculated based on a regression analysis derived from the 15 stimulations (30).

#### 2.4.3 Physiological measurements at baseline and in response to autonomic stress (Figure 1, c-d)

Continuous Heart Rate (HR) and BP at rest and in response to autonomic stress were measured over five minutes using a CareTaker device (https://caretakermedical.net/en-gb/home-2/), placed on the ring fingers of the right hand. Cold pain stimulation, used to induce autonomic stress, was delivered via a locally-developed aluminium probe (dimensions: 4 cm wide x 20 cm long), attached to the volar surface of the left forearm, through which cold water at 4°C was constantly circulated by means of two chillers (Figure 2; see also (31) for a similar procedure). During these recordings, participants were placed in a relaxed, supine position and instructed not to move.

**Figure 2.**
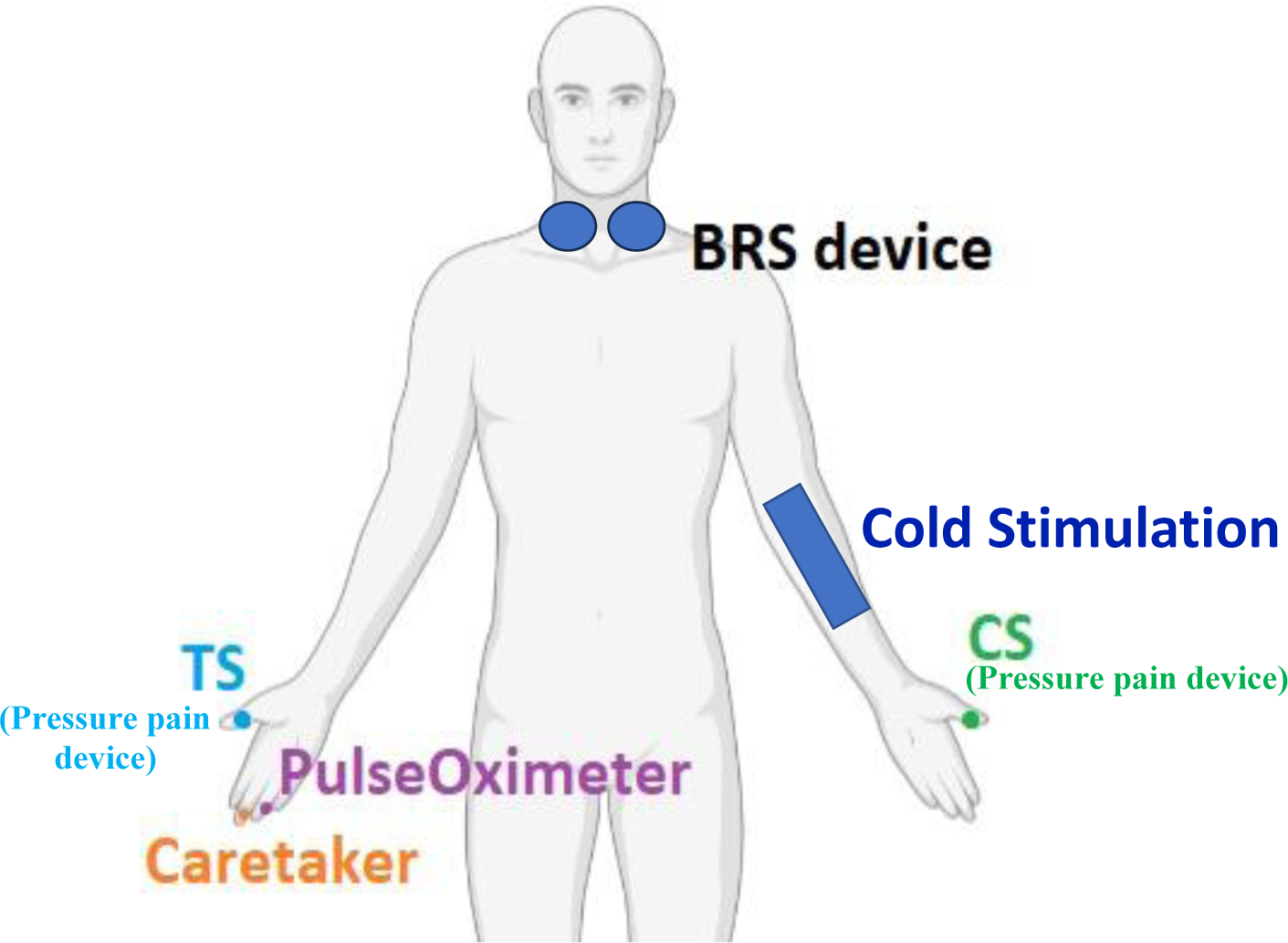
Graphic representation of the experimental set-up. Participants remained connected to the neck suction device throughout the duration of the experiment. The activation of the neck suction (baroreceptor stimulation) was synchronised with the presentation of pain stimuli in all experimental paradigms. Cold stimulation via an aluminium probe was used during the autonomic stress only and removed from participant’s arm after the stimulation, to avoid carry-over effects. Pressure pain (*test stimulus-TS*) was delivered to the right thumbnail, whereas the *conditioning stimulus* (CS) was delivered to the left thumbnail. Continuous HR and BP were collected from the middle and ring fingers, respectively.

#### 2.4.4 Experimental paradigm assessing the effect of *baroreceptor stimulation* on Physiology (Figure 1, e)

Participant’s HR was measured during both the ACTIVE and SHAM conditions to assess the impact of baroreceptor stimulation without any painful stimulation. To examine the physiological response to baroreceptor stimulation, a Pulse Oximeter was used. The baroreceptor stimulation lasted for 8 seconds, followed by a 12-second inter-trial interval, as per the durations used in previous studies (26, 27), which are known to cause activations in the autonomic nervous system and subsequent baroreceptor recovery. A total of 8 ACTIVE and 8 SHAM stimulations were delivered, for a total duration of the experimental task of 8 minutes.

#### 2.4.5 Experimental paradigm assessing the effect of *baroreceptor stimulation* on Pressure pain (Figure 1, f)

PRESSURE PAIN task; Here we examined the effect of the activation of baroreceptors on the perception of pressure pain, delivered simultaneously with either ACTIVE or SHAM artificial baroreceptor stimulation. Negative neck suction started 6 seconds before the painful stimuli, to facilitate baroreceptor activation (Figure 3). Pressure pain (2 second duration) was delivered to the right-hand thumbnail, at the end of the stimulation interval. The total duration of baroreceptor stimulation was 8 seconds. Each pressure pain stimulus was followed by a 12-second rating interval, which also allowed for the recovery of the baroreceptors. Each trial lasted for 20 seconds. Overall, participants rated 8 Pressure + ACTIVE and 8 Pressure + SHAM trials, for a total duration of the experimental run of 5 minutes and 33 seconds.

**Figure 3.**
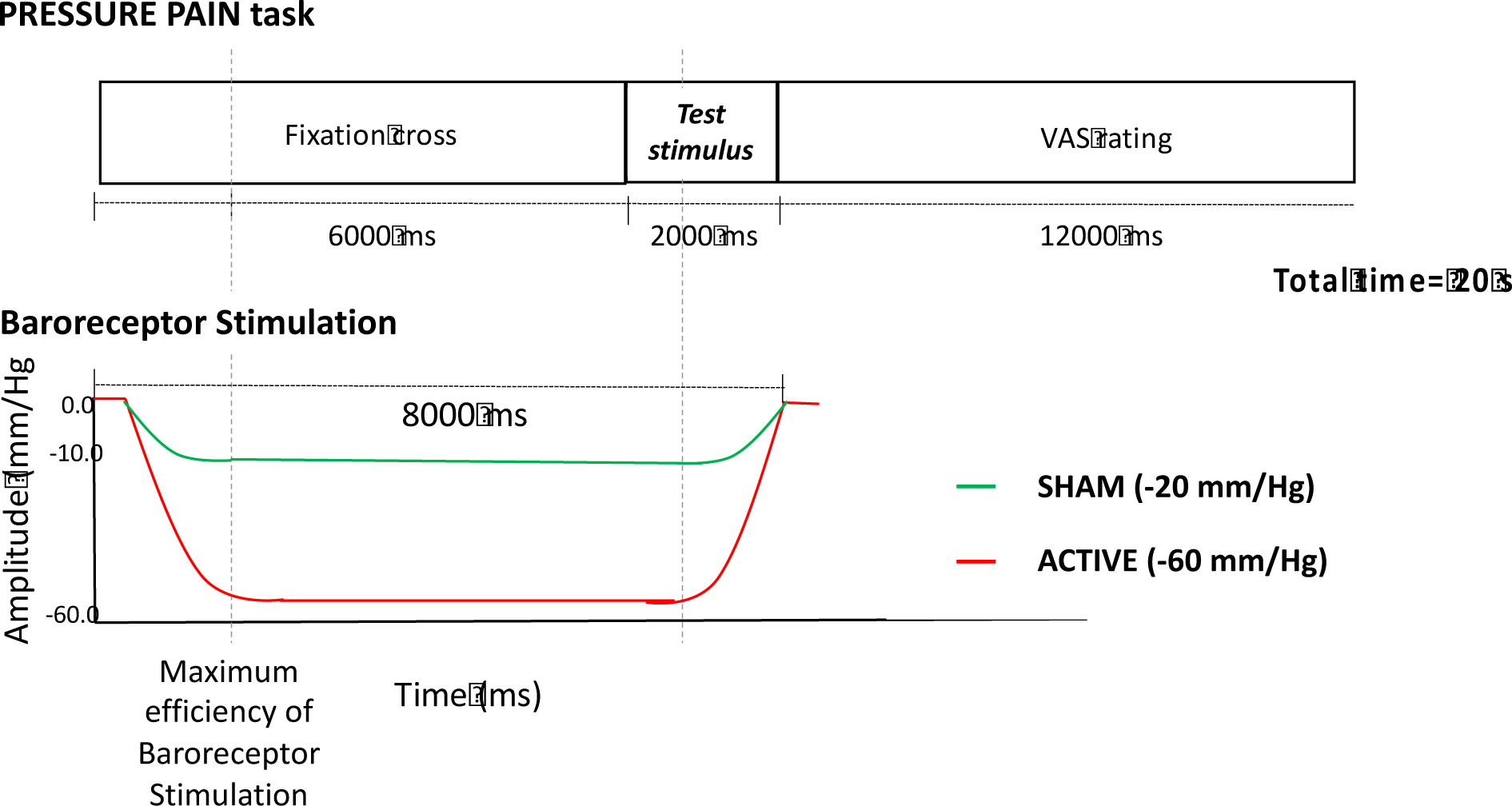
Graphical representation of one experimental trial during the Pressure pain paradigm. Pressure pain was administered for 2 seconds on the right-hand thumbnail, either during ACTIVE or SHAM baroreceptor stimulation. Negative neck suction was initiated 6 seconds prior to the painful stimuli to activate the baroreceptors. Following each pressure pain stimulation, a 12-second rating (VAS) interval allowed for baroreceptor recovery.

#### 2.4.5. Experimental paradigm assessing the effect of *baroreceptor stimulation* on CPM (Figure 1, g)

*CPM task;* Here participants were presented with two types of trials: ‘Pressure only’ and ‘CPM’. During ‘Pressure only’ trials, pressure pain was used as a *test stimulus*, delivered to the right-hand thumbnail. Each trial started with a fixation cross of 2000 ms duration (Figure 4). Participants then received one *test stimulus* (2000 ms duration), followed by a VAS rating period (11000 ms duration) during which participants rated the intensity of their pain on a scale from 0 to 100.

**Figure 4.**
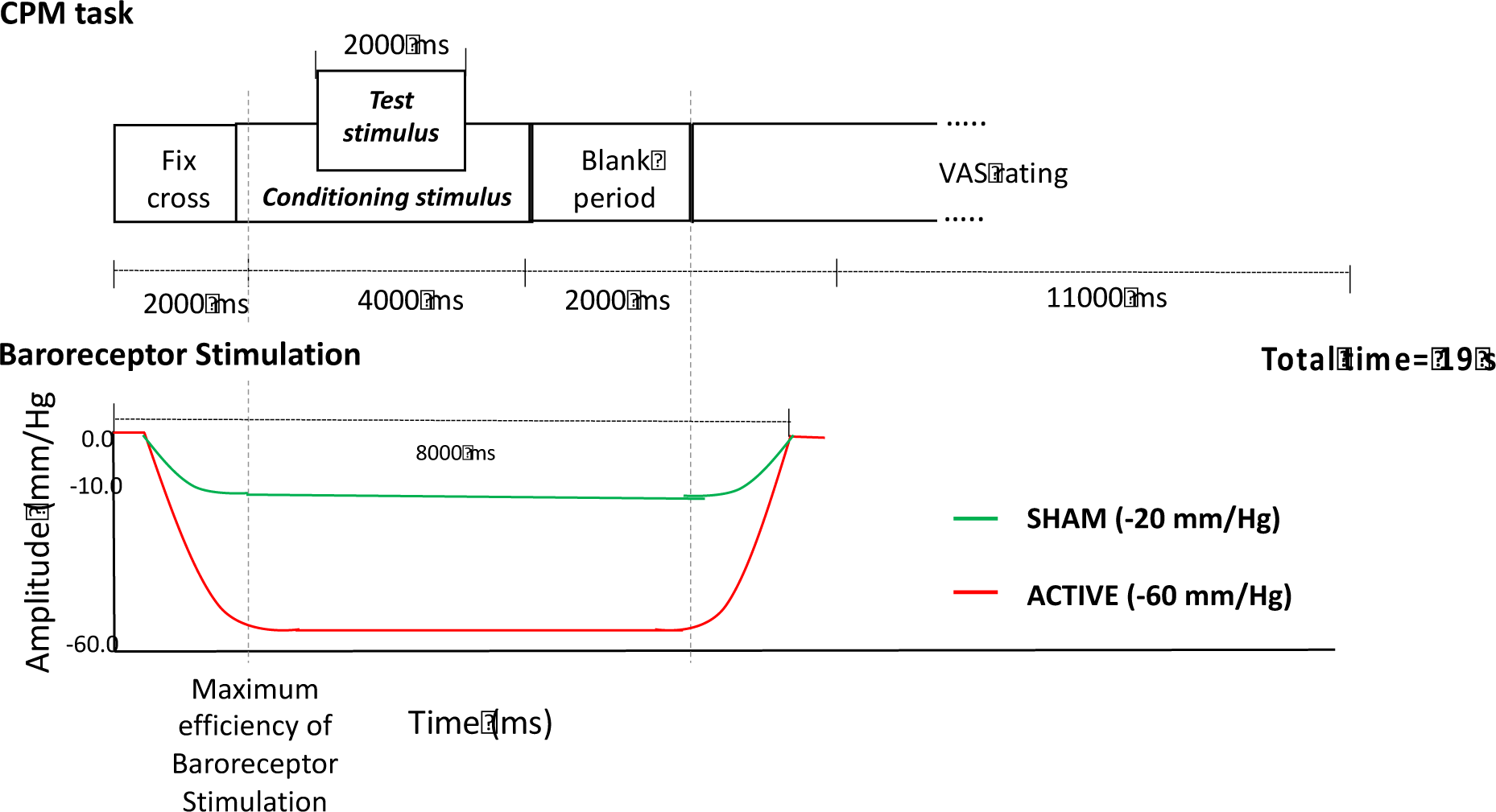
Graphical representation of one experimental trial during the CPM paradigm. During ‘CPM’ trials, participants received a pressure pain *test stimulus* either on its own (‘Pressure only’ trials), or concurrently with a *conditioning stimulus* (CPM trials). Both trials involved random delivery of painful stimulations accompanied by either ACTIVE or SHAM baroreceptor stimulation.

During ‘CPM’ trials, the same *test stimulus* was repeated with a simultaneous *conditioning stimulus,* which consisted in a continuous pressure stimulation (4000 ms duration) applied to the left thumbnail. Participants were instructed to rate the perceived pain in response to the *test stimulus* and to ignore the pain elicited by the *conditioning stimulus*. During both the ‘Pressure only’ and ‘CPM’ trials, painful stimulations were delivered with a simultaneous ACTIVE or SHAM baroreceptor stimulation, which were randomly delivered. The baroreceptor stimulation reached its maximum negative pressure at the onset of the *conditioning stimulus*. The *test stimulus* was delivered 3000 ms following the onset of the baroreceptor stimulation, and 1000 ms following the onset of the *conditioning stimulus* (Figure 4). Each trial lasted 19 seconds. Participants completed a total of 8 trials: 2 ‘Pressure only’ trials and 2 ‘CPM’ trials, each with ACTIVE or SHAM baroreceptor stimulation, randomly delivered. The task lasted 2.53 minutes.

### 2.5 Statistical analysis

Normality of the variables was examined using Shapiro-Wilks tests. Non-normally distributed variables were logarithmically transformed before proceeding with further analyses. All data are expressed as means and standard deviation. Parametric tests were used when normality was achieved; otherwise, non-parametric tests were selected. Age and gender were included as covariates in all analyses. Significance was determined at p < 0.05. Data analyses were conducted using SPSS 23.0 for Windows (SPSS Inc., United States).

#### 2.5.1 Pre-processing of physiological data

Inter-beat (RR) intervals were identified for data collected by means of the CareTaker and PulseOximeter. CareTaker RRs were automatically detected by the CareTaker software. PulseOximeter inter-beat-intervals (IBIs) were calculated with an in-house Matlab 9.7 (R2019b) script (https://www.mathworks.com/products/matlab.html) running on a Linux platform. RR values were visually inspected, and potential artifacts removed manually. RRs were then further corrected by using a threshold-based artefact correction algorithm, adapted from the software Kubios HRV Standard ver. 3.0.2 (32). The algorithm compared each IBI to a local median calculated from 5 nearby heartbeats. If the IBI deviated from the local median beyond a specified threshold, it was considered an artifact and replaced with the median value. Different threshold values (0.45, 0.35, 0.25, 0.15, or 0.05 seconds) were used in a sequential manner, from conservative to liberal. The appropriate correction threshold was selected based on the severity of the individual artifact and followed the procedure described in the Kubios manual (https://www.kubios.com/hrv-preprocessing/). Firstly, we identified beat intervals requiring corrections. If present, we selected the minimum correction level, rectifying the abnormal beat without excessively altering the remaining data.

#### 2.5.2 Baroreflex Sensitivity Index

The Baroreflex Sensitivity Index (BSI) is an index of baroreflex responsiveness, indicating the degree of control of the baroreflex over HR in response to changing BP levels (33). The BSI was calculated from HR and BP data collected during 5-minute rest and cold conditions, using the sequence method, described in (34). Briefly, the method is based on the identification of three or more consecutive beats in which progressive increases/decreases in systolic blood pressure are followed by progressive lengthening/shortening of RR intervals. The threshold values for including beat-to-beat systolic blood pressure and RR interval changes in a sequence were set at 1 mmHg and 6 ms, respectively. The slope of the linear regression line fitted to each sequence was calculated, representing the BSI. The average slope value was used as a measure of BSI.

#### 2.5.3 Analysis of the autonomic activity during autonomic stress

The effect of autonomic stress (cold pain) on BSI and RR values was evaluated by means of a 2X2 ANOVA, with Cold (ON, OFF) and Group (HC, CLBP) as main factors. Age and gender were used as covariates of no interest.

#### 2.5.4 Analysis of autonomic activity in response to *baroreceptor stimulation*

To investigate the effect of baroreceptor stimulation on physiology, mean RR values were calculated from 8-second time windows, corresponding to the baroreceptor stimulation window. The effect of baroreceptor stimulation on RR intervals was investigated by means of a 2X2 ANOVA, with Condition (ACTIVE, SHAM) as the main within-subject factor, and Group (CLBP, HC) as a between-subjects factor.

We then derived the RR_active score (see also Table 1), defined as the difference in RR interval between the ACTIVE and SHAM condition (Active – Sham). Here, negative numbers indicated a decrease in the RR interval during ACTIVE in comparison to SHAM conditions. RR_active scores were used in correlational analyses to explore the association between the physiological reaction to baroreceptor stimulation and behavioural variables (Table 1).

**Table 1.** Description of physiological and behavioural measures.

| Name | Description | Unit |
| --- | --- | --- |
| ***Physiological measures*** |  |  |
| **BSI_rest** | Baroreceptor Sensitivity Index and HR at rest, in supine position | mean slope |
| **HR_rest** |  | ms |
| **BSI_stress** | Baroreceptor Sensitivity Index and HR during autonomic stress (cold pain), in supine position | mean slope |
| **HR_stress** |  | ms |
| **HR_active** | HR during artificial baroreceptor stimulation, in supine position | ms |
| ***Behavioural pain measures*** |  |  |
| **PRESSURE_active** | The effect of ACTIVE or SHAM baroreceptor stimulation on pressure pain perception | VAS |
| **PRESSURE_sham** |  | VAS |
| **Delta_PRESSURE** | The difference in pressure pain perception derived by subtracting the rating of pressure pain in SHAM – ACTIVE condition | Delta VAS |
| **CPM_active** | The activity of the descending pain modulation during ACTIVE or SHAM baroreceptor stimulation, defined as the difference in subjective pain rating between the 'Pressure only' and the 'CPM' condition | Delta VAS |
| **CPM_sham** |  | Delta VAS |

\*Due to technical difficulties, continuous BP was not measured during *baroreceptor stimulation*, thus not allowing for calculations of the BSI\_active index

#### 2.5.5 Analysis of behavioural data from PRESSURE PAIN paradigm

In the PRESSURE PAIN task, the effect of baroreceptor stimulation on pressure pain was investigated with a 2X2 ANOVA, with Condition (ACTIVE, SHAM) as the main within-subject factor, and Group (CLBP, HC) as between-subject factor.

Next, differential scores were calculated for pain intensity ratings, to facilitate correlational analyses (Table 1-Behavioural measures). Delta_PRESSURE was calculated as the difference in VAS pain ratings between the SHAM and ACTIVE conditions. Here, negative values indicated a decrease in perceived pain intensity during the ACTIVE baroreceptor stimulation in comparison to the SHAM stimulation, whereas positive values indicated an increase in perceived pain intensity during the same condition.

#### 2.5.6 Analysis of behavioural data from the CPM paradigm

In the CPM paradigm, we adopted a 2×2×2 ANOVA to explore the effect of the within-subject factors Pain condition (‘Pressure only’, ‘CPM’) and baroreceptor stimulation (ACTIVE, SHAM), and the between-subject factor Group (CLBP, HC).

Next, differential CPM scores were calculated separately for the ACTIVE and SHAM conditions, with the following formula: *CPM_active/sham= VAS (‘CPM’) – VAS (‘Pressure only’)* (Table 1). Here, negative values indicate a decrease in perceived pressure pain during the CPM condition, whereas positive numbers indicate an increase in perceived pressure pain.

#### 2.5.7 Correlational analyses

We investigated the association between physiology measures (systolic and diastolic BP, BSI_rest/HR_rest, BSI_stress/HR_stress, HR_active), clinical scores (Pain State and Trait) and our behavioural measures indicating the VAS rating during painful stimulation (CPM_active, CPM_sham, PRESSURE_active, PRESSURE_sham, Delta_PRESSURE) (See Table 1). We computed correlation analyses, namely, Pearson’s r (for normally distributed data) or Spearman’s test (when normality of the data was not achieved by means of log transformation). Correlations were computed within both the whole sample of HC and patients (to identify associations which were present independently of the chronic pain condition), and within the two groups separately, to seek different patterns of associations in HC and CLBP patients.

## 3 Results

### 3.1 Sample characteristics

Participants’ demographic and clinical characteristics are detailed in Table 2. CLBP patients and HC did not differ in their pressure pain thresholds in either left or right hand (Table 3). CLBP patients’ levels of pain during the past year and on the day of testing are reported in Table 3. Most patients reported weekly pain episodes, which interfered with their work and sleep (Table 3). We did not observe significant differences in standing or sitting BP values between patients and HC (supine systolic BP, HC= 120.6 +/− 9.7, CLBP patients= 119.4 +/− 7.7, p=0.6; supine diastolic BP, HC= 69.1 +/− 7.9, CLBP patients= 69.3 +/− 9.6, p=0.9).

**Table 2.** Demographical profile of CLBP patients and age and gender-matched HC.

| Demographic Measures |  |  |  |
| --- | --- | --- | --- |
| Measure | HC (n=29) | CLBP (n=22) | *p value* |
| Age (years ± SD) | 33.7 (10.8) | 39.2 (11.7) | $p=0.08$ |
| Gender (% females) | 13/29 | 14/22 | $\chi^2=0.06$, $p=0.8$ |
| Years of Education | 18.25 (2.1) | 18.52 (2.5) | $p=0.9$ |
| BMI | 24.4 (3.5) | 23.6 (3.9) | $p=0.6$ |
| Alcohol consumption (UPW) | 4.1 (4.0) | 5.8 (3.7) | $p=0.1$ |

SD= standard deviation. BMI= Body Mass Index. UPW= Units Per Week.

**Table 3.**
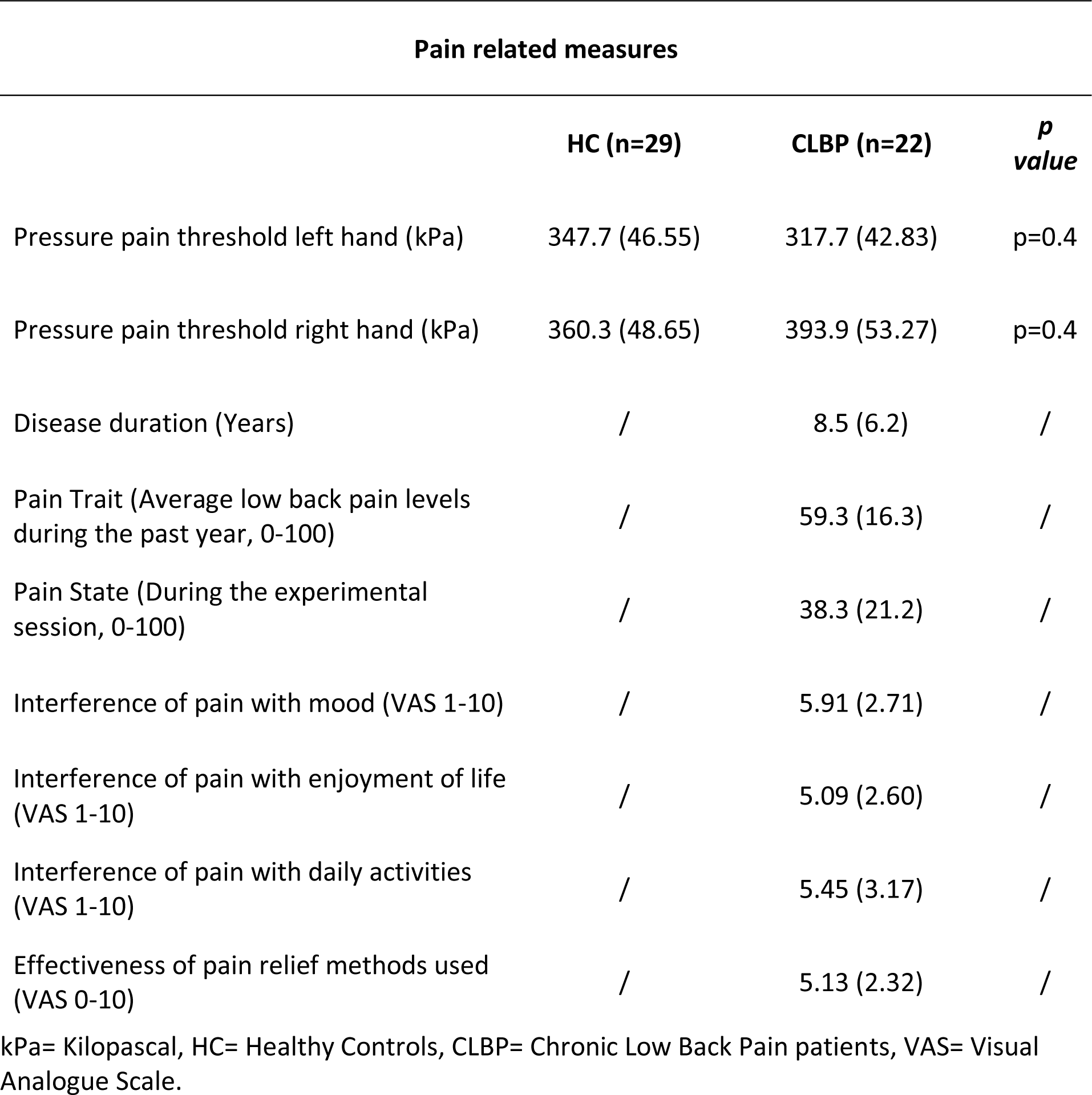
Summary of pain-related measures in the group of HC and CLBP patients.

### 3.2 Effect of autonomic stress on BSI

Two HCs were excluded from the analysis due to a high number of artifacts (>30% of heart beats) in the CareTaker data recorded during cold stimulation. Accordingly, BSI analyses during cold stimulation were performed in 22 CLBP patients and 22 HC. BSI values at rest and during cold were non-normally distributed (W = 0.91, p < 0.002, and W = 0.85, p < 0.01, respectively). Logarithmic transformation was applied to achieve normality. We did not observe a main effect of the factor Cold [F(1,42)=1.0, p=0.32], nor of the Cold x Group interaction [(F(1,42)<1] on BSI., indicating that the autonomic stress of cold stimulation itself did not have a significant effect on BSI, regardless the participant group.

### 3.3 Association between Pain State and Trait in CLBP patients and ANS activity

We observed an association between the clinical levels of Pain Trait and Pain State and BP. Specifically, the Pain State was negatively associated with diastolic BP, where individuals with higher BP reported lower pain on the day of the testing. Pain Trait, similarly, was negatively associated with systolic BP, where individuals with higher BP reported lower levels of pain during the past year (Figure 5).

**Figure 5.**
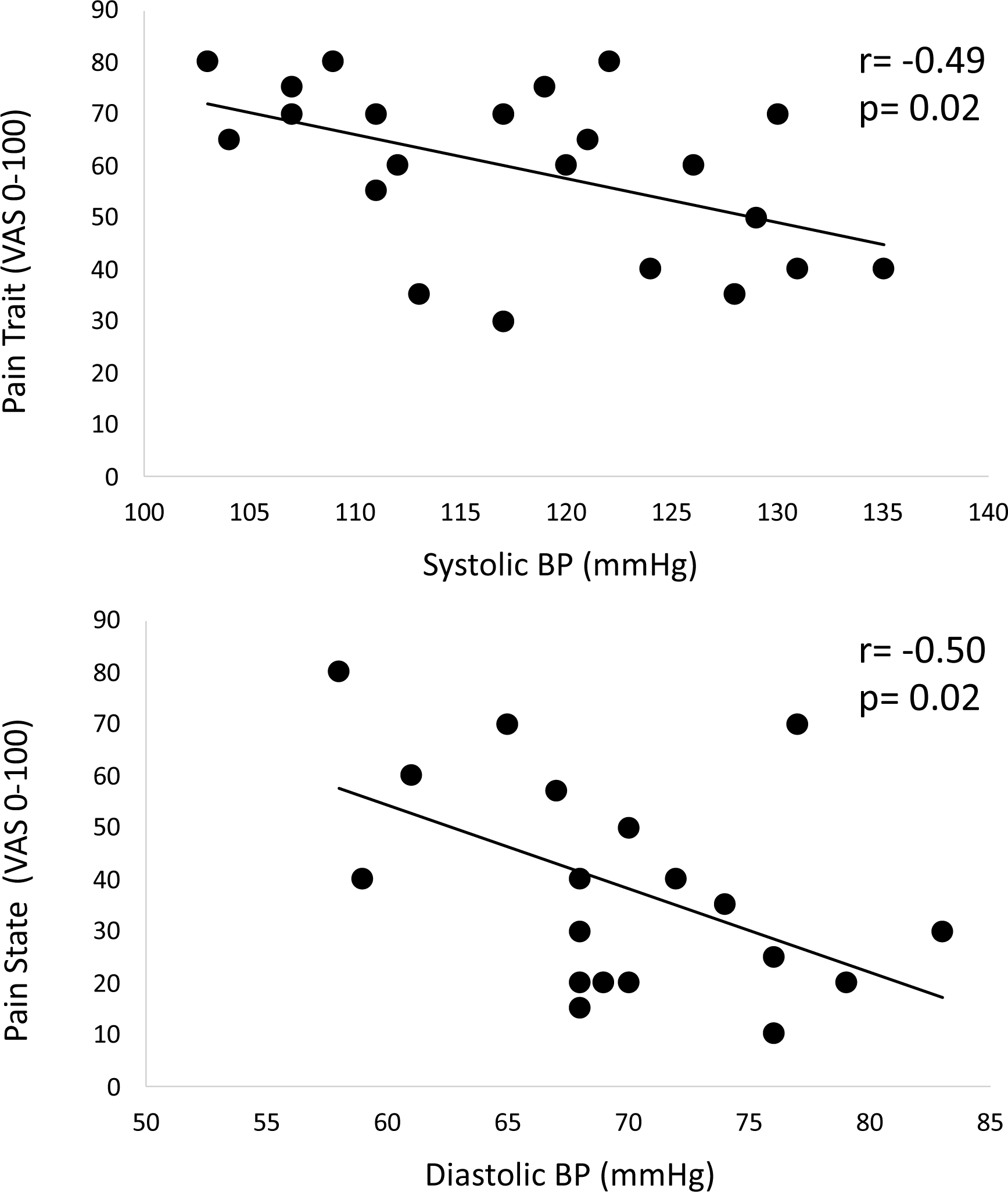
Association between baseline BP and Pain Trait and State measures. A negative association was observed between Pain State and diastolic BP, and Pain Trait and systolic BP.

A positive association was observed between the Pain Trait and BSI_stress, where individuals with higher levels of BSI during cold reported higher levels of low back pain during the past year (Figure 6).

**Figure 6.**
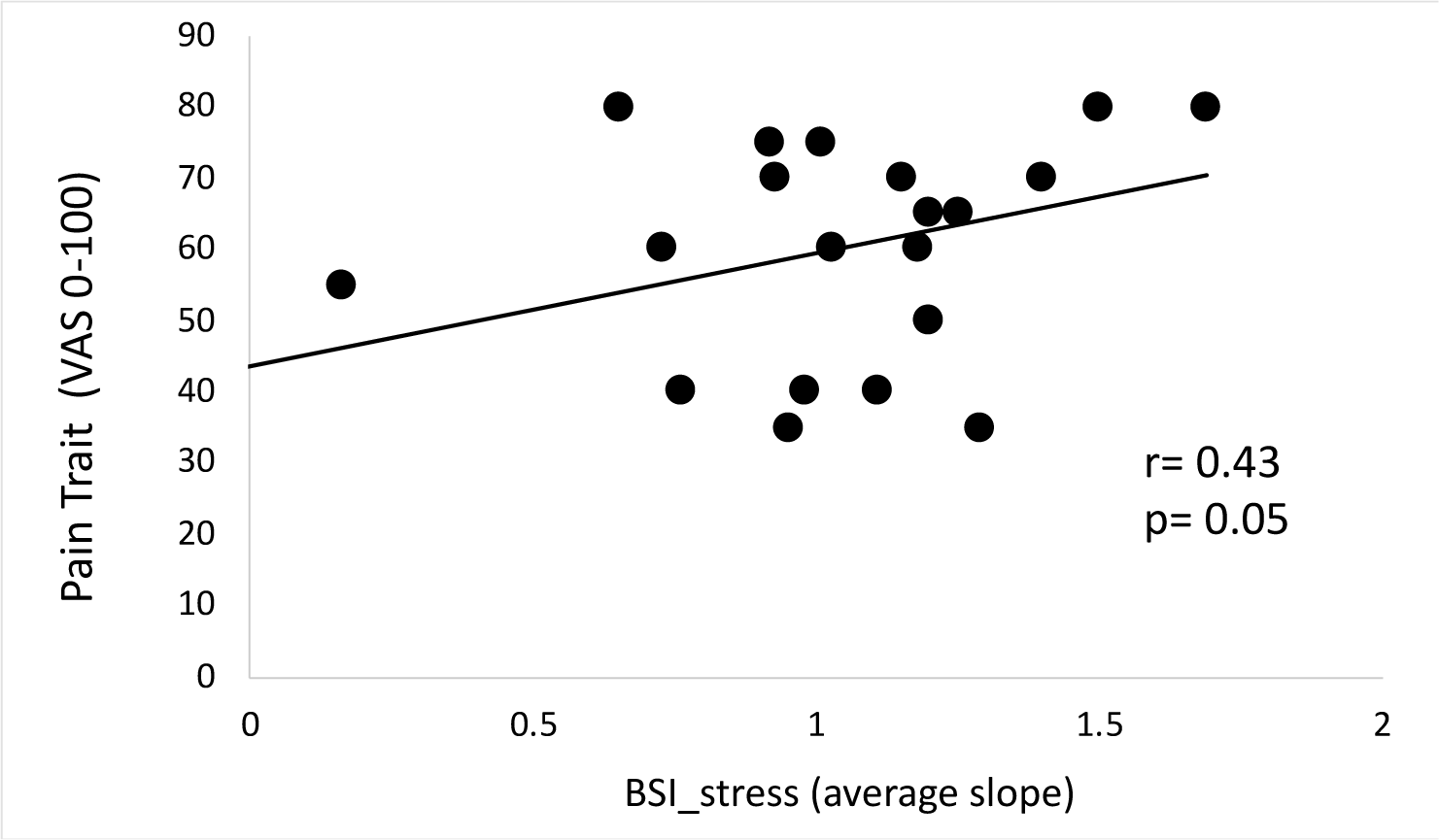
Positive association between Pain Trait and BSI_stress in CLBP patients. Patients with the highest increase in BSI during autonomic stress reported the highest pain during the past year.

### 3.4 Effect of *baroreceptor stimulation* on ANS activity

Overall, when examining the effect of baroreceptor stimulation on IBI, we did not observe a main effect of baroreceptor stimulation (F<1), nor a baroreceptor stimulation x group interaction (F<1). We observed a main effect of the Group [F(1,41)=2.24, p=0.049], driven by lower HR in patients compared to HC [mean and SD patients= 57bpm (+/− 5bpm), HC= 61bpm (+/− 8bpm)].

### 3.5 Effect of *baroreceptor stimulation* on Pressure pain ratings

The distributions were significantly non-normal for the ratings of Pressure pain during ACTIVE (W = 0.94, p = 0.011), and SHAM (W = 0.94, p = 0.013) baroreceptor stimulation, according to Shapiro-Wilk tests. Logarithmic transformation was applied to achieve normality. A 2×2 ANOVA revealed a significant Baroreceptor stimulation x Group interaction [F(1,49)=7.42, p=0.009] (Figure 7).

**Figure 7.**
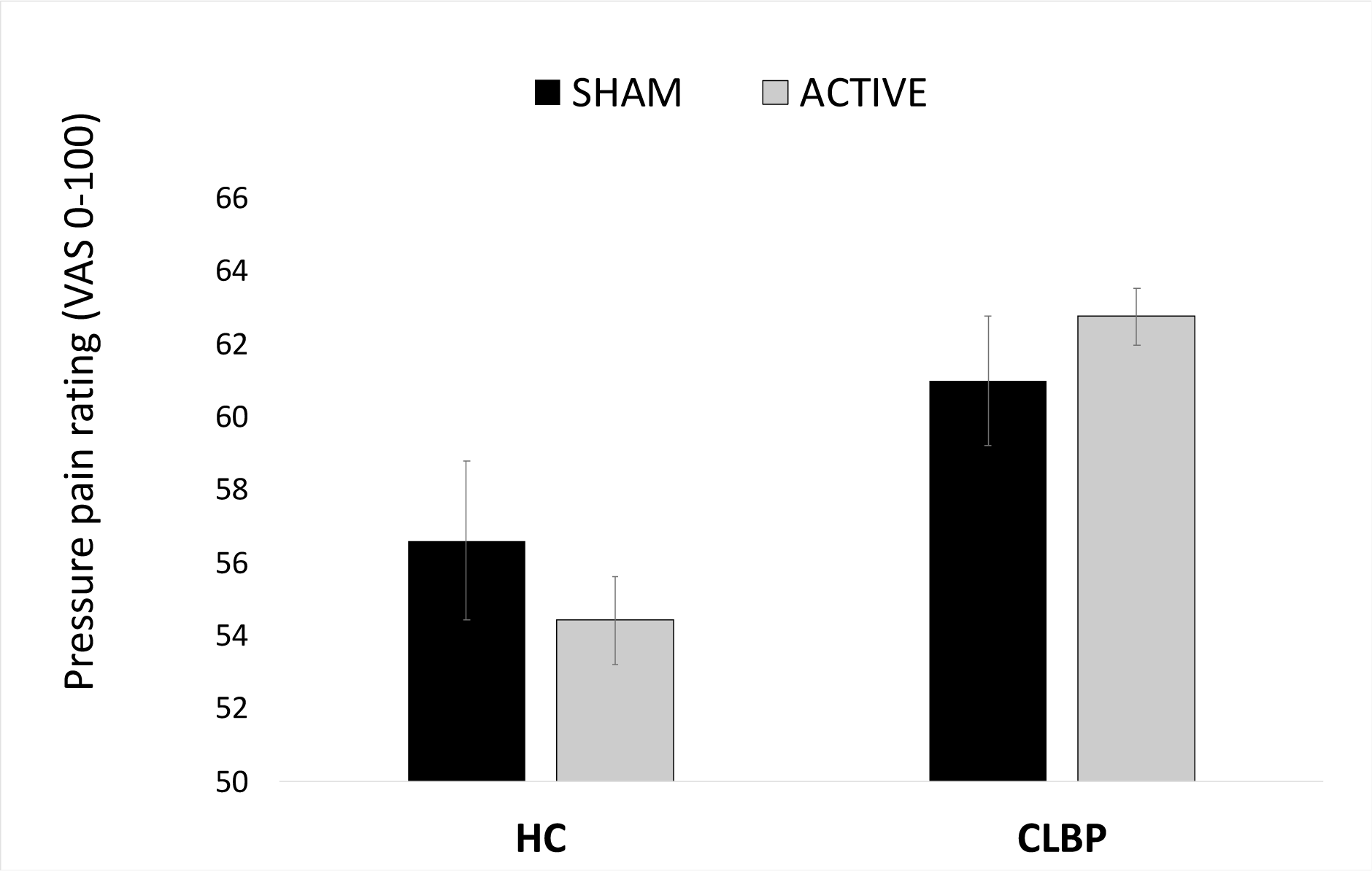
Effect of ACTIVE and SHAM baroreceptor stimulation on perception of pressure pain in HC (n=22) and CLBP patients (n=29). ACTIVE baroreceptor stimulation reduced the perceived pressure pain in HC, and increased pain perception in CLBP patients.

This interaction was driven by a difference in the effect of baroreceptor stimulation on pain perception amongst the two groups of HC and patients, where HC reported a reduction in perceived pressure pain, whereas patients reported an increase in pressure pain (t-test on Delta_PRESSURE, t(49)= 2.6, p=0.013). We did not observe a main effect of Baroreceptor stimulation (F<1) nor for the Group belonging [F(1,49)=2.75, p=0.10].

### 3.6 Effect of baroreceptor stimulation on CPM

We did not observe an effect of baroreceptor stimulation (F<1), nor of the Condition (‘Pressure only’, ‘CPM’) (F<1) or Group (F<1). Similarly, no effect of the baroreceptor stimulation x Condition interaction [F(1,47)= 1.9, p=0.17], of the baroreceptor stimulation x Group [F(1,47)=1.4, p=0.24] or of the Condition x Group interaction (F<1) was observed. Indicating no effect of baroreceptor stimulation on the CPM responses in both groups of HC and CLBP.

### 3.7 Association between BSI_rest and BSI_stress, and PRESSURE PAIN and CPM ratings

In the CPM task, an association was observed across the whole group of HC and CLBP patients between the CPM_active and BSI_rest, indicating that individuals with the highest baroreceptor sensitivity at rest showed the strongest reduction in pain during CPM_active. The same direction of effect was evident during the CPM_sham condition, without reaching statistical significance (Figure 8). No associations were observed between BSI_stress and CPM and Pressure pain scores (all p>0.5).

**Figure 8.**
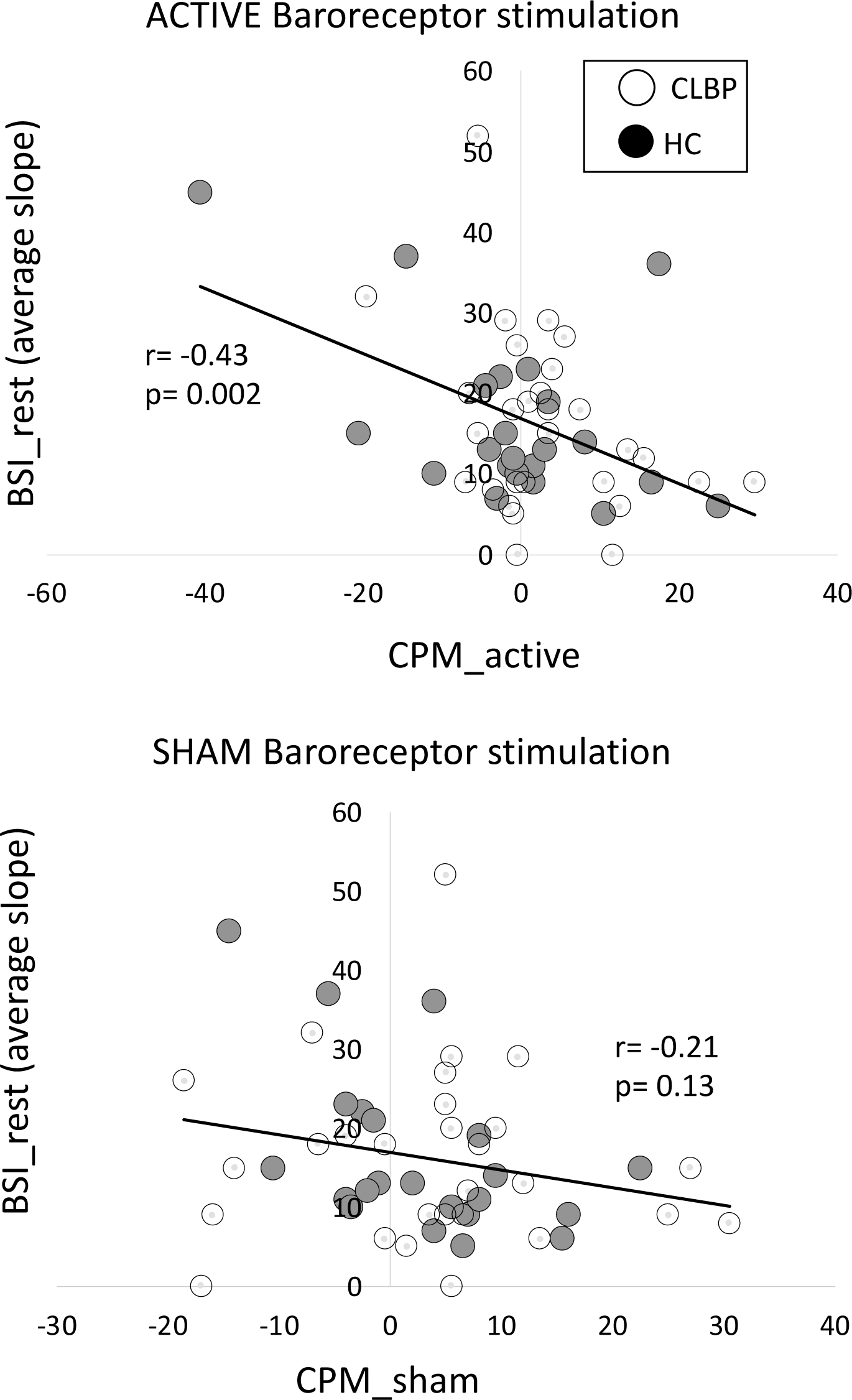
Negative association between BSI_rest and (Upper panel) CPM_active and (Lower panel) CPM_sham across both HC and CLBP patients. Only the association between baroreceptor sensitivity and CPM during ACTIVE baroreceptor stimulation reached statistical significance. BSI_rest= Baroreceptor Sensitivity Index at rest; CPM_active/sham= activity of the descending pain modulation during ACTIVE or SHAM baroreceptor stimulation.

### 3.8 Association between HR_active and PRESSURE PAIN and CPM ratings

We observed a negative association between HR_active changes and the amount of pressure pain modulation induced by ACTIVE *baroreceptor stimulation* during the PRESSURE PAIN task (Figure 9). Here, a decrease in HR in CLBP patients was associated with lower ratings of pressure pain during ACTIVE stimulation (r= −0.46, p= 0.043; Figure 9) in CLBP patients. The same effect was observed in HC; however, it did not reach statistical significance (r= −0.29, p= 0.17). In the CPM task, no association was observed between the HR_active and CPM scores (all p >0.1).

**Figure 9.**
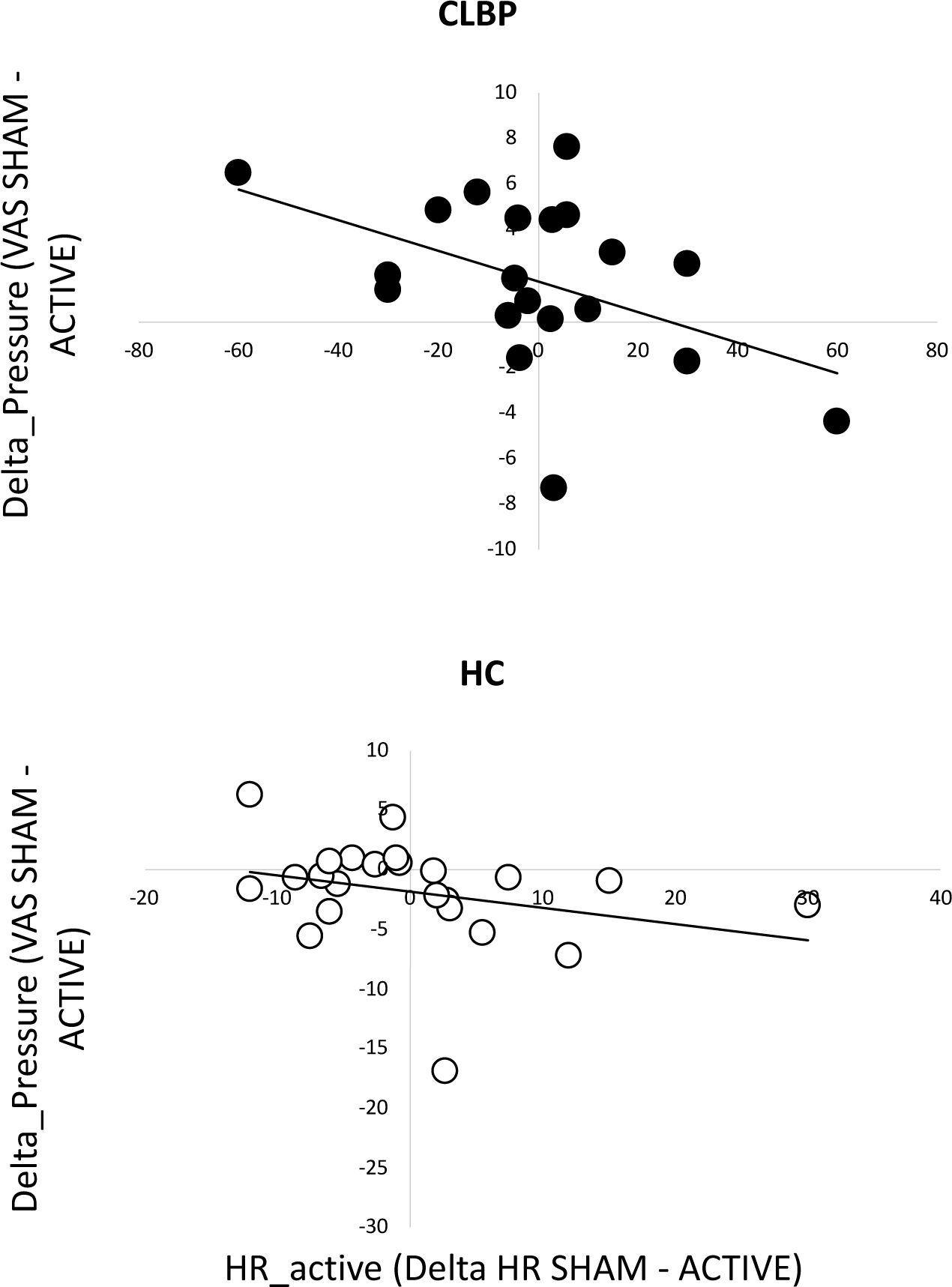
Association between HR_active and pressure pain ratings. A negative association was observed between an increase in HR during ACTIVE baroreflex stimulation and a decrease in pressure pain ratings in patients (Upper panel). The same effect was evident in HC; however, it did not reach statistical significance (Lower panel).

## 4 Discussion

ANS activity and BP can influence pain perception (4). Both animal studies (35) and experimental studies with manipulation of baroreceptors in humans (17) suggest that the baroreflex accounts at least in part for cardiovascular–pain association. While the nociceptive and the autonomic nervous systems interact at peripheral, spinal cord, brainstem, and forebrain levels, the mechanisms underlying this relationship are not fully described. In this study, we investigated the association between the baroreflex, pain from a noxious pressure stimulus, and descending pain modulation, in a group of healthy controls and patients with CLBP. We used artificial baroreceptor stimulation to explore whether the baroreflex differentially modulates pain perception in HC and CLBP patients. In our patients, autonomic activity at rest, as indicated by resting BP, showed a consistent association between BP and pain experience of patients. However, when the baroreflex was stimulated, via baroreceptor stimulation or autonomic stress, the association between the baroreflex and pain involved distinct mechanisms in patients with chronic pain and healthy controls, indicating that the reactivity of the baroreflex might be compromised in chronic pain conditions.

We further identified a negative association between BP and measures of low back pain (both State and Trait pain) in patients, supporting theories indicating that higher BP levels may be protective against pain. Higher diastolic BP was associated with reduced back pain during the testing session (Pain State). Similarly, higher systolic BP was associated with lower levels of pain during the past year (Pain Trait). Finding this relationship intact in patients partially disagrees with previous studies that suggested that the relationship between BP, thermal pain thresholds, and pain tolerance is disrupted in chronic pain (36). However, pain threshold and tolerance in response to acute experimental pain might not adequately capture the mechanisms of pain-ANS association in patients who are in ongoing pain. Alternatively, a preserved association between BP and clinical pain may be due to the otherwise relatively healthy cardiovascular system of patients in our cohort, as only patients without ANS dysregulation (as assessed, for example, by the Valsalva manoeuvre) were selected. Close control of cardiovascular risk may have precluded the inclusion of patients in whom an association between BP, specifically, baroreflex efficiency, and descending pain control may be compromised. Future studies should strive to enhance our understanding of these mechanisms by incorporating chronic pain patients with elevated cardiovascular risk. Additionally, it is crucial to investigate the specific stage in the chronicity of pain where the association between blood pressure and pain deviates or becomes disrupted.

Further analyses revealed an alteration in the association between the baroreflex and pain in CLBP patients, in conditions where the baroreflex was activated either by autonomic stress or by artificial baroreceptor stimulation. The reactivity of the baroreflex during autonomic stress demonstrated a positive correlation with Pain Trait scores, indicating that individuals with the highest increase in BSI during autonomic stress reported the most severe pain experienced over the past year. In healthy individuals, physiological (i.e. cold stimulation) or psychological stress is known to reduce BSI (37), possibly due to inhibitory control originating from the hypothalamus and targeting the nucleus tractus solitarius (38). Similarly, another study by Sabharwal et al. (2004) found that cold exposure reduced baroreflex sensitivity in rats (39). This reduction is likely due to activation of the sympathetic nervous system, which can cause vasoconstriction and an increase in blood pressure. Therefore, transient reductions in BSI during stress is a normal reaction and likely part of a well-functioning homeostatic mechanisms. Previous studies have shown that individuals with chronic pain exhibit blunted cardiovascular reactivity to stress (40) and orthostatic challenges (41), indicating that the inhibitory effect of stress on the baroreflex might be dysregulated in these patients. Consequently, the influence of the baroreflex on descending pain modulation pathways may shift towards pain facilitation rather than pain inhibition during stress. Our results are in line with this hypothesis. Indeed, baroreflex stimulation reduced perceived pressure pain in HC, whereas the opposite pattern was observed in CLBP patients. Further, a positive correlation was observed in HC between the impact of baroreflex stimulation on pain and its effect on physiological responses. On the other hand, this relationship was disrupted in patients, aligning with previous studies involving chronic pain patients.

Contrary to our initial hypotheses, we did not observe differential effects of baroreceptor stimulation on the CPM response between HC and patients with CLBP. A significant association was, however, observed between BSI_rest and the magnitude of inhibition/facilitation during CPM in HC and CLBP. Both patients, and HC with lower resting BSI levels exhibited facilitation, that is, an increase in subjective pain perception during CPM. This finding mirrors our previous study, where the CPM response in HC was associated with low frequency heart rate variability (LF-HRV) index (42), a measure associated with the baroreflex efficiency (43, 44). Replicating this finding in an independent group of individuals, validates our initial hypotheses of an association between the baroreflex and descending pain control capacity. MRI studies have shown that brainstem regions important in both invoking CPM responses and elaboration of afferent cardiovascular information, namely the PAG and rostral ventrolateral medulla, are also implicated in the aetiology and maintenance of chronic pain (45, 46). Further MRI analyses in our previously published study with HC revealed that the association between LF-HRV and CPM was mediated by the strength of functional connectivity a statistical measure of the association of activity between anatomically distant brain regions) between the periaqueductal grey and the prefrontal cortex (42). Currently, untangling whether dysregulation of the relationship between autonomic responses and descending pain responses is a result of living with chronic pain or a predisposing factor remains challenging. Longitudinal data are required to provide answers to this question. However, the existence of this association in HC within our study suggests the presence of a potential vulnerability that may be found in non-clinical populations. We speculate that such vulnerability may contribute to the development of chronic pain and that the successful characterisation of this mechanism may be of benefit for identifying patients at risk of developing chronic pain.

This study indicates the potential importance of the baroreflex in pain perception and indicates potential avenues of exploiting these mechanisms to improve treatment of patients with chronic pain. Improved understanding of the mechanisms underlying these findings may facilitate provision of individualised pain management strategies. Pharmacological or non-pharmacological treatments targeting the baroreflex might also be explored as potential interventions for chronic pain. For example, further exploring the role of the alpha-2 adrenergic system in stimulating baroreflex sensitivity may provide to novel pharmacological targets for chronic pain. While the precise mechanisms remain unknown, dexmedetomidine, an α2-adrenergic receptor agonist with proven baroreflex modulation (47), has demonstrated analgesic properties (48). This observation further reinforces the hypothesis of the central role of the baroreflex in pain modulation. Additionally, non-pharmacological interventions such as slow breathing, stress reduction techniques, and heart rate variability (HRV) biofeedback show promise in alleviating chronic pain (49, 50). Stratifying patients based on underlying pathophysiological mechanisms could help identify those who are more likely to benefit from interventions targeting the baroreflex. Ultimately, these advancements have the potential to enhance pain management approaches and improve the quality of life for individuals with chronic pain.

We acknowledge some methodological limitations in our study. For practical reasons, we collected heart rate data using a photoplethysmography (PPG) rather than using electrocardiographic (ECG) recordings, the gold standard approach for deriving HRV measures. Some studies have shown that under certain conditions, PPG can serve as a suitable alternative of ECG-derived HRV (51, 52). We are aware of the debates around the optimal CPM paradigm, however, the types of painful stimulation adopted in this study, namely, pressure-pain delivered to the thumbnail bed as a test stimulus, and cold-pain as a conditioning stimulus have been previously reported as approaches with good reliability (21).

## 5 Conclusions

Our data provide evidence that indicates the importance of baroreflex functioning in pain modulatory processes in HC and patients and provide a first experimental attempt to elucidate the role of the baroreceptor in descending pain modulation in patients with chronic pain. The findings are important for the validation of baroreflex-associated autonomic measures of pain vulnerability and maintenance in chronic pain syndromes. We suggest they provide enticing potential as markers of pain severity and movement towards much-needed mechanism-based profiling of patients with chronic pain.

## Data Availability

All data produced in the present study are available upon reasonable request to the authors.

## Acknowledgments

We thank all volunteers for their participation in the study and the Brixton Therapy Centre for their help with recruitment. We also express our gratitude to Dr. Giovanni Calcagnini for his contribution to the development of the baroreceptor stimulating machine, and extend our appreciation to Alfonso De Lara-Rubio and Simon Hill for the realisation of the associated hardware and software.

## Notes

### Competing Interest Statement

The authors have declared no competing interest.

### Funding Statement

This study was funded by the Effic-Grunenthal Award 2019 and the Medical Research Council. Dr Matthew Howard is supported by the NIHR Biomedical Research Centre and Clinical Research Facility at South London and Maudsley NHS Foundation Trust and King's College London.

### Author Declarations

Research Ethics committee of King's College London gave ethical approval for this work.

